# Mutation induced infection waves in diseases like COVID-19

**DOI:** 10.1101/2021.07.06.21260067

**Authors:** Fabian Jan Schwarzendahl, Jens Grauer, Benno Liebchen, Hartmut Löwen

**Affiliations:** Institut für Theoretische Physik II: Weiche Materie, Heinrich-Heine-Universität Düsseldorf, 40225 Düsseldorf, Germany; Institute of Condensed Matter Physics, Technische Universität Darmstadt, Darmstadt, Germany

## Abstract

After almost 4 million deaths worldwide, the ongoing vaccination to conquer the COVID-19 disease is now competing with the emergence of increasingly contagious mutations, repeatedly supplanting earlier strains. Following the near-absence of historical examples of the long-time evolution of infectious diseases under similar circumstances, models are crucial to exemplify possible scenarios. Accordingly, in the present work we systematically generalize the popular susceptible-infected-recovered model to account for mutations leading to repeatedly occurring new strains, which we coarse grain based on tools from statistical mechanics to derive a model predicting the most likely outcomes. The model predicts that mutations can induce a super-exponential growth of infection numbers at early times, which can self-amplify to giant infection waves which are caused by a positive feedback loop between infection numbers and mutations and lead to a simultaneous infection of the majority of the population. At later stages – if vaccination progresses too slowly – mutations can interrupt an ongoing decrease of infection numbers and can cause infection revivals which can occur as single waves or even as whole wave trains featuring alternative periods of decreasing and increasing infection numbers. Our results might be useful for discussions regarding the importance of a release of vaccine-patents to reduce the risk of mutation-induced infection revivals but also to coordinate the release of measures following a downwards trend of infection numbers.

## Introduction

The COVID-19 pandemic [1, 2] has led to more than 150 million infected [3] and more than 3.4 million death [3] worldwide until the end of Mai 2021. During the course of the pandemic the SARS-CoV-2 virus has mutated into various different strains [4, 5], some of which have led to an increased infection rate [6–8] as compared to the original strain [2] (Wuhan 2019). Examples are the variants B.1.1.7 and B.1.351, which have driven a strong rise of infection numbers in the United Kingdom and South Africa [9, 10] in late 2020 [11, 12] and the P.1 mutation which has induced an infection wave in Brazil [13] in early 2021.

The availability and ongoing vaccine production gives hope to slowly gain control of the disease [14–16]. However, before herd immunity is finally reached worldwide it will take many month or even years, which the virus will exploit to mutate into a range of new strains. Thus, at the timescale of months or years a race is looming ahead between the occurrence of new mutations and the adaption and mass-production of existing vaccines to get these mutations under control – making it questionable if the present vaccination program is sufficiently effective to ultimately get COVID-19 under control. It is therefore important to understand possible mutation-induced long-time disease-evolution scenarios e.g. in view of the ongoing discussions regarding the release of patents to accelerate worldwide vaccination but also regarding requirement of measures like social distancing once the infection numbers show a downwards trend.

Notably, historical examples to assess possible longtime consequences of mutation cascades are scarce, since particularly severe mutations have traditionally led to a rapid death of infected individuals eliminating these mutations. Thanks to modern medical treatment based e.g. on extracorporeal membrane oxygenation support or artificial aspiration, however, such a self-elimination of severe mutations is largely absent. Notably, besides the positive effect of immediately saving many lives, these treatments also have the side effect of inducing a potentially disastrous self-amplification of mutations and infection-rates. This hinges on the facts that (i) mutations can serve as seeds for further mutations which are even more infectious than the strain from which they have emerged; (ii) mutation rates are higher when infection numbers are high. Both factors together can generally lead to a positive feedback loop between infection numbers and mutations suggesting severe long-time mutation-induced effects for the disease evolution. Actual data for COVID-19 mutations show, in fact, early signatures supporting such a possible self-amplification scenario: They reveal an initial constant and a subsequent nonlinear growth of the relevant infection rate (Fig. 1(a)). Thus, it would be highly important to understand the possible long-time consequences of such a self-amplification mechanism and how fast vaccination has to progress worldwide in order to suppress the most dramatic ones. However, following the scarcity of useful historical examples illustrating the possible consequences, we have to rely on models to explore the possible impact of mutations on the long-time evolution of the disease dynamics, in particular also in the presence of vaccination and other actions counteracting the self amplification mechanism. To provide a concrete starting point for such an exploration, in the present work, we develop a statistical minimal model to predict possible mutation-induced effects for the long-time evolution of infectious diseases like COVID-19. We first develop a stochastic multi-strain generalization of the popular susceptible-infected-recovered (SIR) model to account for the random occurrence of mutations and then use the coarse-graining concept of statistical physics to derive an effective mean-field model enabling general predictions of the most likely scenarios for a given scenario (characterized by parameters such as the mutation and the vaccination rate). See Fig. 1 (b) for an schematic illustration of our approach.

**FIG. 1.**
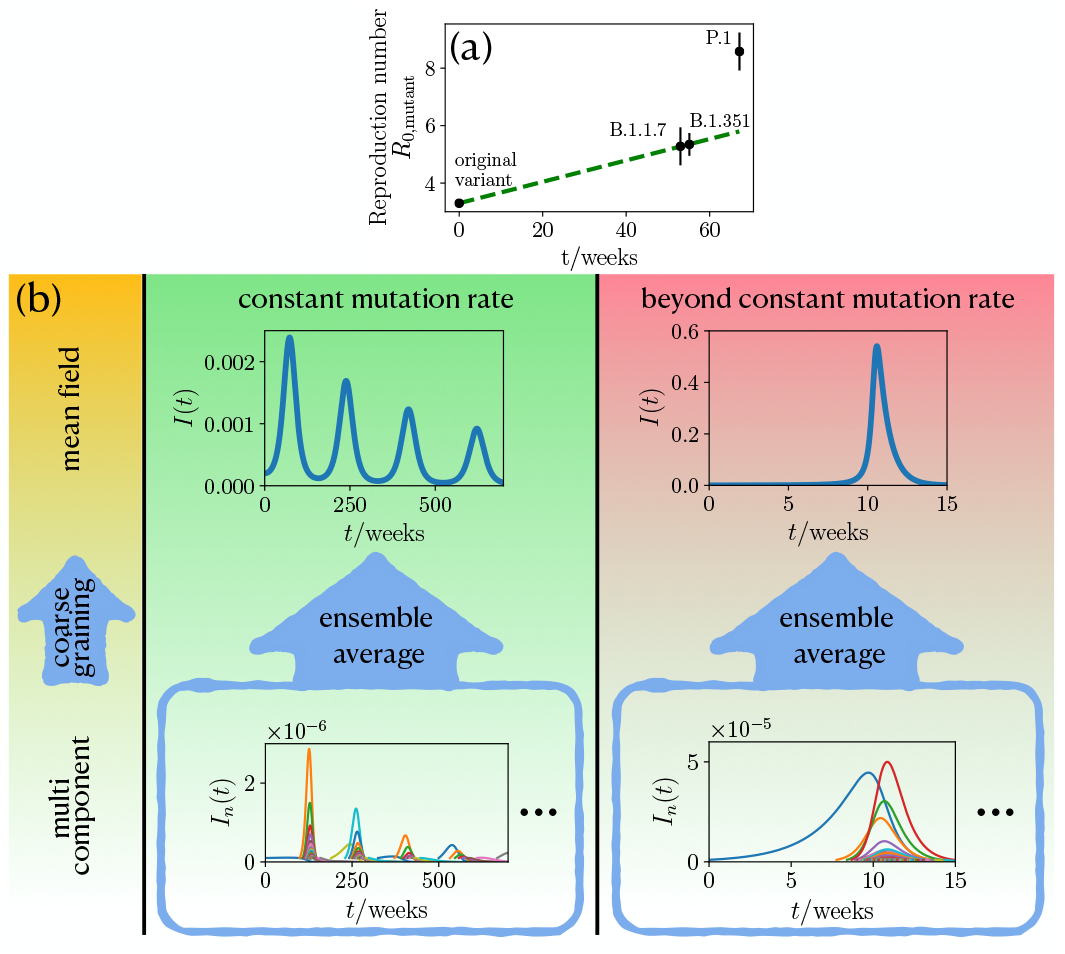
The statistics of mutation formation and its impact on the course of an epidemic. (a): Reproduction number for different COVID-19 mutations as a function of the their emergence time. (for details see Methods) Green dashed line shows a linear fit to initial constant growth. (b): For a constant mutation rate (middle panel, green background) an ensemble average of the multi component description with many infection strains *I*_*n*_(*t*) leads to multiple infection waves of the global infection number *I*(*t*). Beyond the constant mutation rate, if the mutation rate is coupled to the infection number (right panel, red background), the ensemble average produces a hidden singularity, as manifested by the giant infection wave. Only one representative realization of the multi component description is shown in the blue frames. The left panel (yellow background) indicates the different levels of description starting from multi components leading to an effect mean field by coarse graining.

One generic prediction of our model is that mutations induce an *explosive super-exponential growth* of the infection numbers rather than the ordinary and much discussed normal exponential growth, in phases where the population is far away from herd immunity. At later phases, when a population comes close enough to herd-immunity that the reproduction number drops below one (*R <* 1) and infection numbers subsequently decrease to a very low level, mutations can raise the reproduction number to *R >* 1 inducing a new infection wave, which is followed by a whole train of further waves. This scenario occurs even for a constant mutation rate (Fig. 1 (b)). If the mutation rate increases with the number of infections, as generally expected and discussed above, their effect is even more dramatic: then, mutations occur at a self-accelerating pace and continuously prevent the population from reaching herd-immunity by persistently enhancing the effective reproduction number of the disease. As a result the infection dynamics approaches a hidden singularity and displays signatures of a critical dynamics. That is, infection numbers grow extremely fast, giving a giant infection wave, such that the majority of the population is infected at the same time (see the values on the vertical axis in Fig. 1 (b)), which would massively overstrain any existing medical system. Finally, in phases where vaccination of the population takes place and is sufficiently effective to suppress the hidden singularity and hence the explosive self-acceleration of infection numbers, our model predicts the possibility of mutation-induced infection wave trains, as in the case of constant vaccination, illustrating once more the possible dramatic consequences following from the fact that herd-immunity is not necessarily a permanent state in the presence of mutations. To see how these predictions come about, let us now discuss our modelling approach in detail.

## Model

To describe the impact of mutations on the infection dynamics within a simple statistical framework, we first generalize the popular susceptible-infected-recovered (SIR) model [17–29], which has been intensively explored in the context of the COVID-19 pandemic [30–41]. While some recent works have generalized this model to account for two different infectious strains [42, 43], here we allow for the continuous emergence of new strains with a rate *ν*, which in general depends on the present infection number. Denoting the fraction of susceptible and recovered individuals with *S* and *R* respectively and the fraction of individuals which is infected with strain *n* as *I*_*n*_, this leads use to the following dynamical equations:

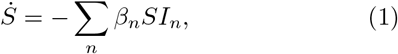

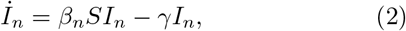

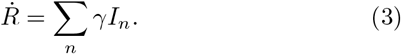

Here, *γ* is the inverse of the average disease duration, i.e. the recovery/death rate and *β*_*n*_ is the infection rate of strain *n*, which we randomly choose from a certain characteristic distribution. As an initial state, we assume that initially (time *t* = 0) we have only a single infectious strain with a low positive infection number such that only *I*_0_ ≳ 0 whereas *I*_*n*=0_ = 0 for *n* = 1, 2…

To allow predicting the average (or most likely) result of the infection dynamics we now coarse grain this model, essentially by averaging over many strains and disease realizations (see Methods section for technical details), which leads to the following effective model:

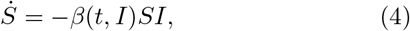

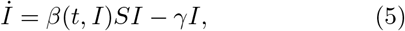

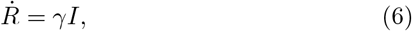

Here *I* is the overall infection number (all strains together) and *β*(*t, I*) is the average infection rate, which can depend on the overall infection number, depending on the underlying mutation statistics (see Methods).

## Results

Let us now explore the impact of mutations on the disease evolution by comparing numerical simulations of the multi-component model with analytical predictions based on the mean-field model (Fig. 1 (b)). To allow distinguishing between direct effects of mutations from the prevailing (and most infectious) strain and indirect effects due to the self-accelerating mutation cascade which we have described in the introduction and which may or may not become effective in reality, depending on the actual mutation rate and other parameters, we will sequentially follow on the cases of (i) a constant mutation rate *µ* leading to the emergence of new strains in our simulations with a constant rate and (ii) a mutation rate which depends on the present infection number *µ*(*I*).

### Constant mutation rate

#### Mutation-driven infection dynamics

Let us assume that the infection rate *β*_*n*_ of a newly occurring virus-strain is randomly selected from a normal distribution *p* with standard deviation *σ* centered around the infection rate of the presently prevailing strain:

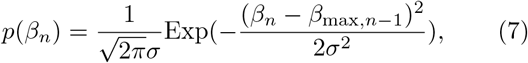

Here *β*_max,*n-*1_ denotes the largest infection rate of all currently existing strains. This distribution, the average of which moves towards higher values in the course of a disease, is motivated by the fact that newly occurring mutations typically become visible only if they have a higher (or at least not much lower) infection rate than the currently prevailing ones. Coarse graining this mutation statistic (see Methods) yields an the following average infection rate for our mean-field model

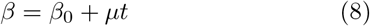

where *µ* is the constant mutation rate and *β*_0_ is the initial infection rate. That is, coarse graining the distribution (7) leads to a constant increase of the infection rate with time.

Let us first explore the disease evolution at early times when the majority of the population is susceptible, such that *S*(*t*) ≈ 1. Then Eq. (5) reduces to

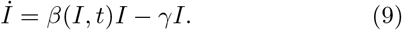

Now using Eq. (8) we find

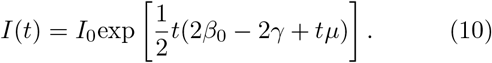

Thus, the fraction of infected individuals does not grow exponentially as in the standard SIR model but even faster. Following Eq. (10) if the mutation rate is high enough that *tµ* ≫ 2(*β*_0_ *γ*) long before herd-immunity is reached, the infection dynamics generically converges towards 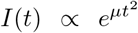, which is completely mutation driven. To test this prediction, we now numerically solve the full multi-component model and show the overall *I*(*t*) = ∑_*n*_ *I*_*n*_(*t*) in Fig. 2(a). Notably, the result is close to the analytical prediction of the mean-field model and shows an even slightly larger growth.

**FIG. 2.**
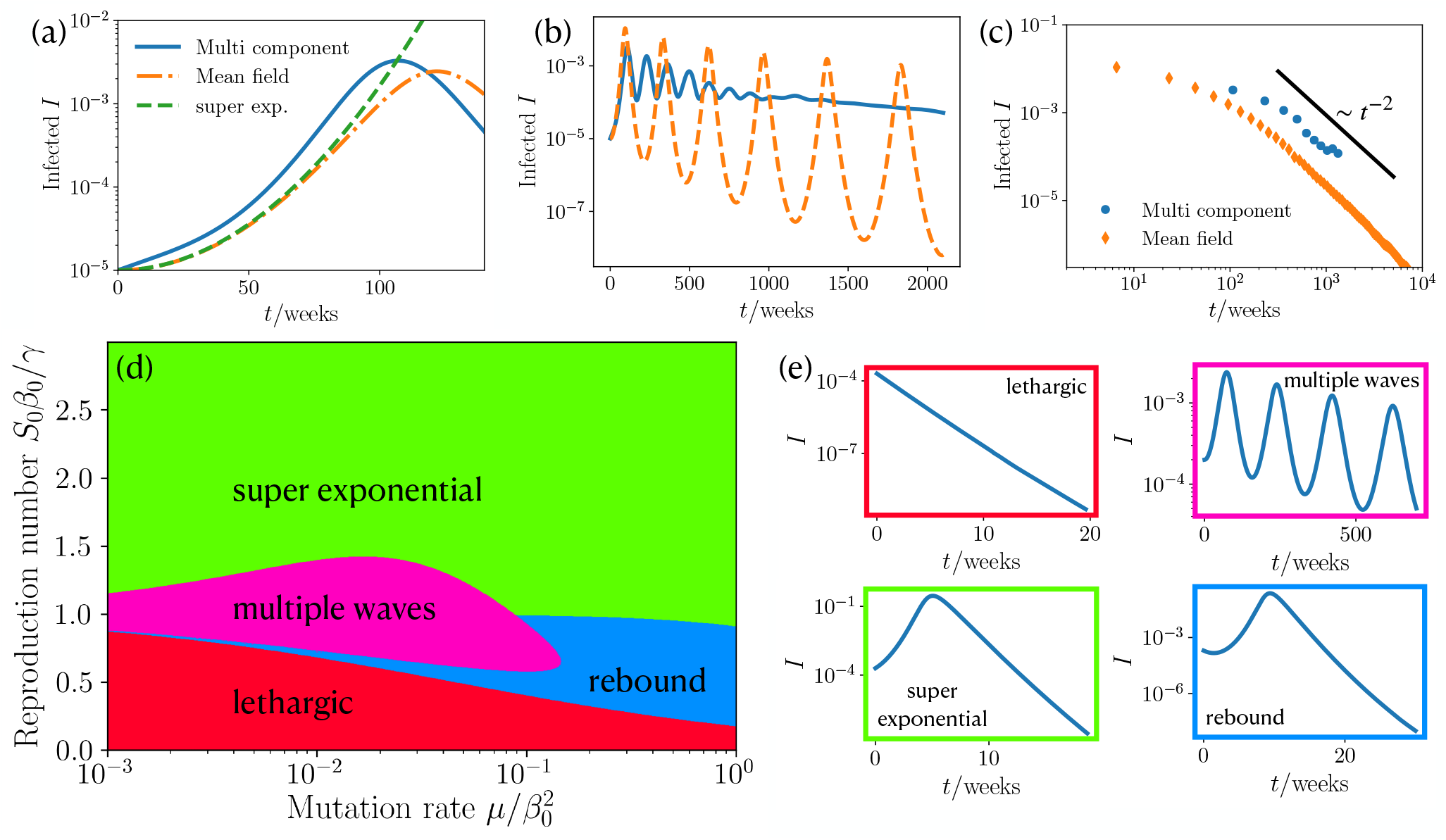
Power law dependence of infection dynamics, phase diagram and state classification for constant mutation rate. (a) Fraction of infected people *I* at short times *t* for the coarse grained MSIR, multi component MSIR and the early time approximation Eq. (10). (*I*_0_ = 10^−5^, *R*_0_ = 1, 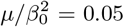) (b) Long time wave pattern of fraction of infected people for coarse grained MSIR and multi component MSIR approach. (*I*_0_ = 10^−5^, *R*_0_ = 1, 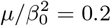) (c) Scaling of maxima of infections for coarse grained MSIR and multi component MSIR approach. (*I*_0_ = 10^−5^, *R*_0_ = 1, 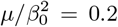) (d) Phase diagram for our coarse grained infection dynamics showing the occurrence of four different courses of the pandemic for varying mutation rate and reproduction number. (*I*_0_ = 2 × 10^−4^) (e) Different courses of infections during an epidemic: lethargic, multiple waves, super exponential, and rebound.

Clearly, the predicted (super)exponential growth of the infection numbers can not continue forever but has to saturate once the population reaches herd-immunity either by collectively going through the infection or through vaccination. Once herd-immunity is reached, the infection numbers are normally expected to monotonously decrease, as predicted by the standard SIR model. However, numerical solutions of our mean-field model show that after a phase where the population recovers and infection numbers decay to a very low level, they can rapidly grow again (Fig. 2(b)). This sequence of decreasing and increasing infection numbers can even repeat for many times, leading to an infection wave-train. The maxima of the wave-trains follow a scaling law of *I*_max_ ∼ 1*/*(*β*_0_ + *µt*_max_)^2^ (see SI for derivation), which is shown in Fig. 2(c). The prediction of wave trains is also confirmed by numerical solutions of the multi-component model, but somewhat weakened, because the individual strains can show waves occurring at individual “frequencies”. Let us now ask about the mechanism leading to these infection waves. They are induced by the nonlinear coupling of the infected and susceptible. First, the number of infected people grows, when the term *βS* in Eq. (5) is large enough. At a certain point, the number of susceptible is too small and the saturation effect from the recovery rate *γ* in Eq. (5) takes over such that the number of infected decreases. However, the infection rate *β* continues growing with time, such that *βS* can become large enough to induce a second wave. This feedback continues on multiple times giving rise to the oscillatory behavior shown in Fig. 2(b). One a more intuitive level, these considerations show that in the presence of mutations herd-immunity is not necessarily a persistent state of a population and that strongly decreasing infection numbers are not an overall reliable sign that the population has overcome the disease. From a sociopolitical viewpoint each growth phase within such wave trains might evoke (nonpharmaceutical) inventions, creating an immense mental burden on the population.

#### Phase diagram

To see how strong measures have to be taken to prevent such an infection wave train (or a super-exponential growth) in the first place, we now systematically vary the parameters in the model to create a state diagram providing a systematic overview on the possible scenarios. It turns out that there are three dimensionless control parameters in our system (see SI), one of which is the initial infection number and the other two ones are the effective reproduction number *S*_0_*β*_0_*/γ* and a dimensionless mutation rate 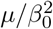. Varying both parameters systematically and solving the meanfield MSIR model for each parameter combination we obtain the phase diagram shown in Fig. 2(e), which shows four qualitatively different epidemic courses: a lethargic phase, which is characterized by an exponential decay, multiple waves, super exponential wave, and a rebound, with an initial local minimum and a proceeding super exponential increase. The occurrence of these states in parameter space is summarized in Fig. 2(d). At large reproduction number the dynamics is “super exponential” (green domain) for any positive mutation rate. When decreasing the reproduction number, depending on the mutation rate, one reaches the regime of “multiple waves” which we have previously discussed (pink) or a “rebound” phase (blue) where infection numbers initially decrease, pass a minimum and then increase to reach a single maximum before finally decreasing (Fig. 2(e)). For even lower mutation rates, the population is in the “lethargic” regime, where the infection numbers monotonously decrease.

#### Nonpharmaceutical inventions and vaccination

In practice the goal is of course be to apply appropriate measures to safely reach the lethargic regime in Fig. 2(d) and not to end up in the multiple wave or rebound regime where the evolution of infection numbers show a promising initial trend but a severe evolution at later times. To understand the impact of nonpharmaceutical inventions, we reduce the reproduction number [30–32, 34, 44, 45], from *R*_0_ to a reduced reproduction number including measures *R*_0,int_ at a time *t*_int_ in our simulations and numerical solutions of the MSIR model. Starting in the super exponential wave regime (*R*_0_ = 1.5) we apply interventions during the rise of a first wave (see Fig. 3(a)). By including weak measures (*R*_0,int_ = 1.3), the maximum of infections decreases as intended, however, the wave needs a longer time to decay, implying a longer period of restrictions for the public. Lowering the reproduction number to *R*_0,int_ = 0.9, results in the appearance of a second and third intervention-mutation induced wave. These waves are enabled by the increased infection rate and the fact that due to the interventions there are more susceptible at a later point in time, where they can facilitate the growth of infections. Here, the situation would be particularly confounding to the public, since it was subjected to measures to decrease the number of infections in the first place; however, this results in more waves and a likely extension of the period of interventions. On the other hand, a strong reduction of the reproduction number (*R*_0,int_ = 0.7) gives a fast decay of infections, as intended. Of course the situation changes when we start from an infection dynamics with multiple waves (*R*_0_ = 1) and apply measures during the rise of the second wave (see Fig. 3(b)). As expected, strong measures (*R*_0,int_ = 0.8) have the intended effect of eliminating the epidemic. On the other hand, if the measures are slightly weaker (*R*_0,int_ = 0.85), the infections first decrease, but then lead to a second intervention-mutation induced delayed wave, which is stronger in magnitude than the first wave. Again, this wave is induced by the growing infection rate and the enhanced number of susceptible individuals due to interventions. To decision makers and the public this type of wave could likely appear as unexpected. However, note that at least the overall number of recovered people at the end of our the epidemic is decreasing with stronger interventions, meaning that if only the cumulative number of infections is considered every reduction of *R*_0_ is useful.

**FIG. 3.**
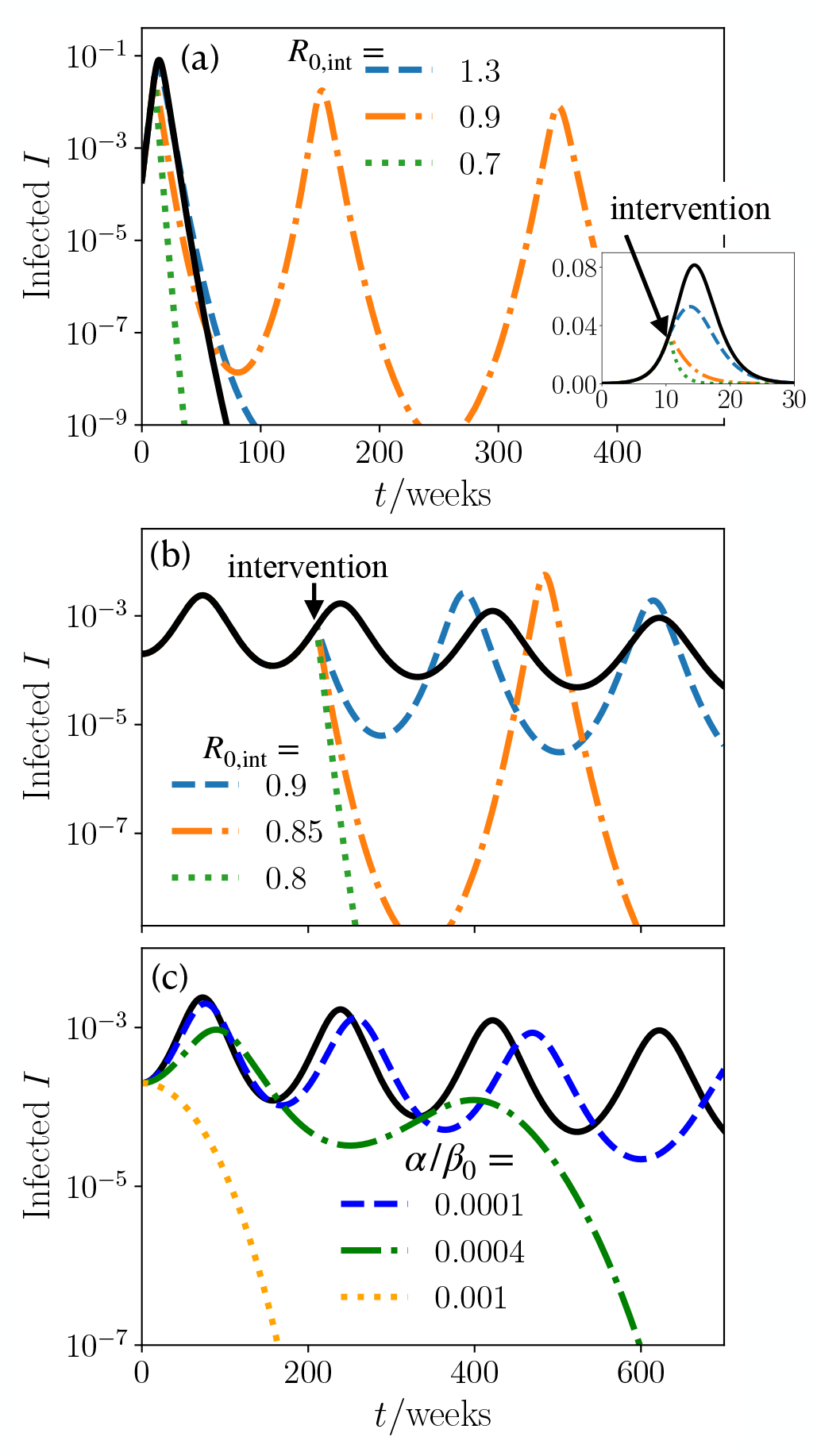
Nonpharmaceutical interventions and vaccinating. (a) Infections as function of time for a super exponential wave. Measures are taken by reducing the reproduction number to *R*_0,int_ at time *t*_int_. Inset: zoom in to the early time regime. (Black line has no interventions, *R*_0_ = 1.5, 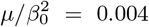, *t*_int_ = 10.5 weeks) (b) Infections as function of time for a wave like pandemic course. Measures are taken by reducing the reproduction number to *R*_0,int_ at time *t*_int_. (Black line has no interventions, *R*_0_ = 1, 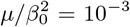, *t*_int_ = 210 weeks) (c) Infections as function of time of different vaccination rates *α*. (Black line has no vaccination, *R*_0_ = 1, 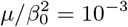, *I*_0_ = 2 × 10^*-*4^)

Specifically for COVID-19 vaccines have recently become available and their continuous production gives hope to overcome the disease soon. However, more than half a year after vaccines have first become available only about 5% of the worldwide population has been fully vaccinated (early June 2021), leaving much time for the emergence of highly infectious mutations. Vaccinations effectively reduce the number of susceptible in an SIR approach [14], such that we modify Eq. (4) as

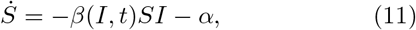

with a vaccination rate *α*. We investigate the effect of vaccination in a infection dynamics with multiple waves (see Fig. 3(c)). Vaccinations have a clear effect, they have to be applied fast enough to significantly reduce the number of infections and temper the train of waves. This further shows the importance of manufacturing and distributing vaccinations as fast as possible.

### Beyond constant mutation rate

Let us now account for the fact that in reality the mutation rate is coupled to the infection number, such that mutations are more likely in phases where the infection numbers are large. To account for this effect in our model, we assume that new (relevant) mutations occur with a probability of *p*_0_*I*_*n−*1_ from the most infectious strain where *p*_0_ is constant and also that the infection rates of new strains follow from a random walk with a mutation-induced bias as *β*_*n*_ = *β*_*n−*1_ + Δ*β* (see Methods for details). Coarse graining the biased random walk yields a mean field infection rate which evolves as

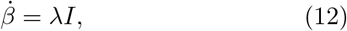

with a mutation rate *λ*. Intuitively, this means that new mutations occur in our mean-field model with a rate which is proportional to the present infection number.

#### Mutation-induced dynamics

At early times, where *S* ≈ 1 we obtain again Eq. (9), which yields together with Eq. (12)

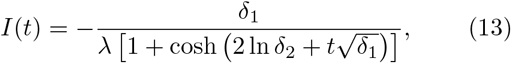

where *δ*_1_ and *δ*_2_ are constants that are given in the SI. Importantly, the infections in Eq. (13) do not grow exponentially, but there is an explosive super exponential growth, which asymptotically has a scaling behavior following *I*_sc_(*t*) ∼ 1*/* |*t* − *t*_*c*_|^2^ with a critical time *t*_*c*_ (see SI for explicit expression). Crucially, this giant infection wave qualitatively differs from the comparatively mild super-exponential behaviour which we have encountered for the case of a constant mutation rate in that it leads to a much more extreme self-acceleration of the infection numbers. As a result, the infection numbers peak only at extremely high values where a large fraction of the population is infected at the same time (see Fig. 1b and Fig. 4(a)). Clearly, such an explosive growth would be interrupted at some point as the population approaches herd immunity. To quantify to which extend the predicted explosive growth would occur before herd-immunity causes significant deviations, we consider the expression min(ln(*I*_sc_) - ln(*I*)), which quantifies how closely the fraction of infections approaches the underlying (idealized) power law dependence in the presence of saturation effects. We find that for large mutation rates the power-law dependence is strong for any reproduction number (Fig. 4(b)) and weakens for lower mutation rates. Remarkably, the explosive growth depends only weakly on the reproduction which is the parameter that is controllable due to interventions. After the initial super-exponential increase of infections the saturation effects from people recovering induce a maximum and a succeeding decrease in infection numbers.

**FIG. 4.**
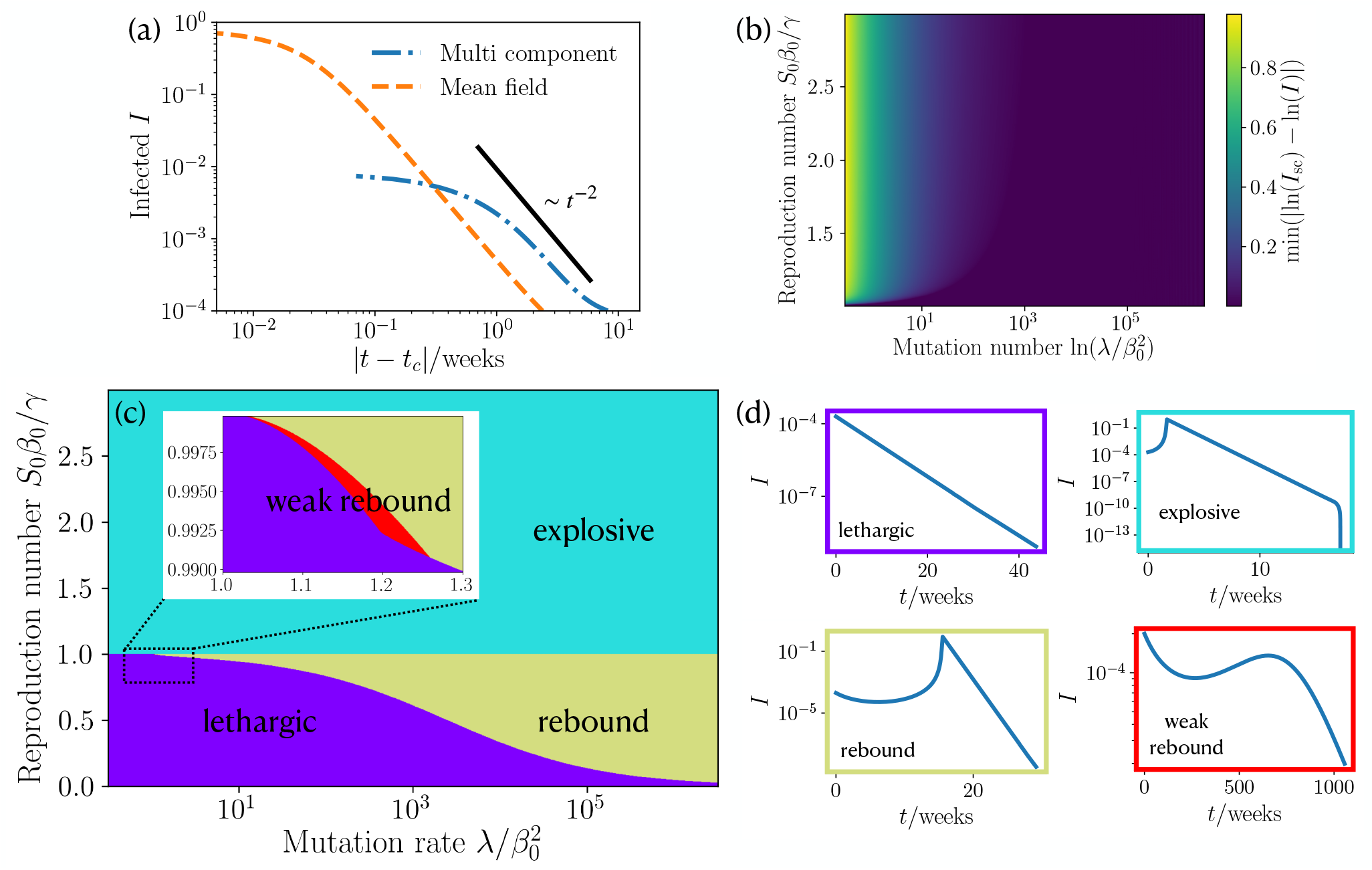
Scaling law of short time infection dynamics, Phase diagram and state classification of approach beyond constant mutation rate. (a) Infections as function of reduced time |*t − t*_*c*_ | where *t*_*c*_ is the critical time at which the infections diverge (see SI for details). We show the scaling law, coarse grained MSIR and our multi component MSIR approach. (*R*_0_ = 2, 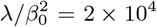, *I*_0_ = 10^−4^) (b) Deviation of the fraction of infections at short times from our corse grained MSIR approach to the a 1*/* |*t − t*_*c*_ | ^2^ scaling by using min(ln(*I*_sc_) − ln(*I*)). Mutation rate and reproduction number are varied. (*I*_0_ = 2 × 10^−4^) (c) Phase diagram of our coarse grained MSIR approach showing the occurrence of four different courses of the pandemic for varying mutation rate and reproduction number. (*I*_0_ = 2 × 10^−4^) (d) Example plots of the infections as function of time for four different courses of the pandemic: lethargic, super exponential wave; rebound, and weak rebound.

#### Phase diagram

Depending on the basic reproduction number and the mutation rate the MSIR model predicts four distinct courses summarized in the phase diagram Fig. 4(c), which has been obtained analytically (see SI). We find a lethargic regime characterized by an exponential decay (purple regime, Fig. 4(d)); an explosive regime (cyan regime); a rebound regime (dark yellow) where we have a minimum followed by a mutant induced super exponential increase; and weak rebound (red) leading to an infection maximum which is smaller than the initial fraction of infected individuals. Generally, the explosive (or super exponential) regime occurs for reproduction numbers *R*_0_ *>* 1 and any positive mutation rate, whereas the other three regimes occur for *R*_0_ *<* 1. For low mutation rate and *R*_0_ *<* 1 the epidemic is in the desired lethargic regime, increasing the mutation rate leads to a small region of weak rebound which then transitions to a rebound dynamics.

#### Nonpharmaceutical inventions and vaccination

To explore the efficiency of measures which effectively reduce the reproduction number, we again change the reproduction number *R*_0_ to a value of *R*_0,int_ at time *t*_int_. Now starting from the explosive (super exponential) regime (*R*_0_ = 1.2) we decrease the reproduction number during the rise of the wave (Fig. 5). Strong measures yielding *R*_0,int_ = 0.7 or *R*_0,int_ = 0.8) induce an immediate decay of the infection numbers as desired. However, weak measures, leading to *R*_0,int_ = 1 only delay the occurrence of the infection maximum but hardly change the value of *I* at the peak. Hence, it is clear that measures need to be strong enough to have a significant effect, while weak measures only delay the infection number explosion.

**FIG. 5.**
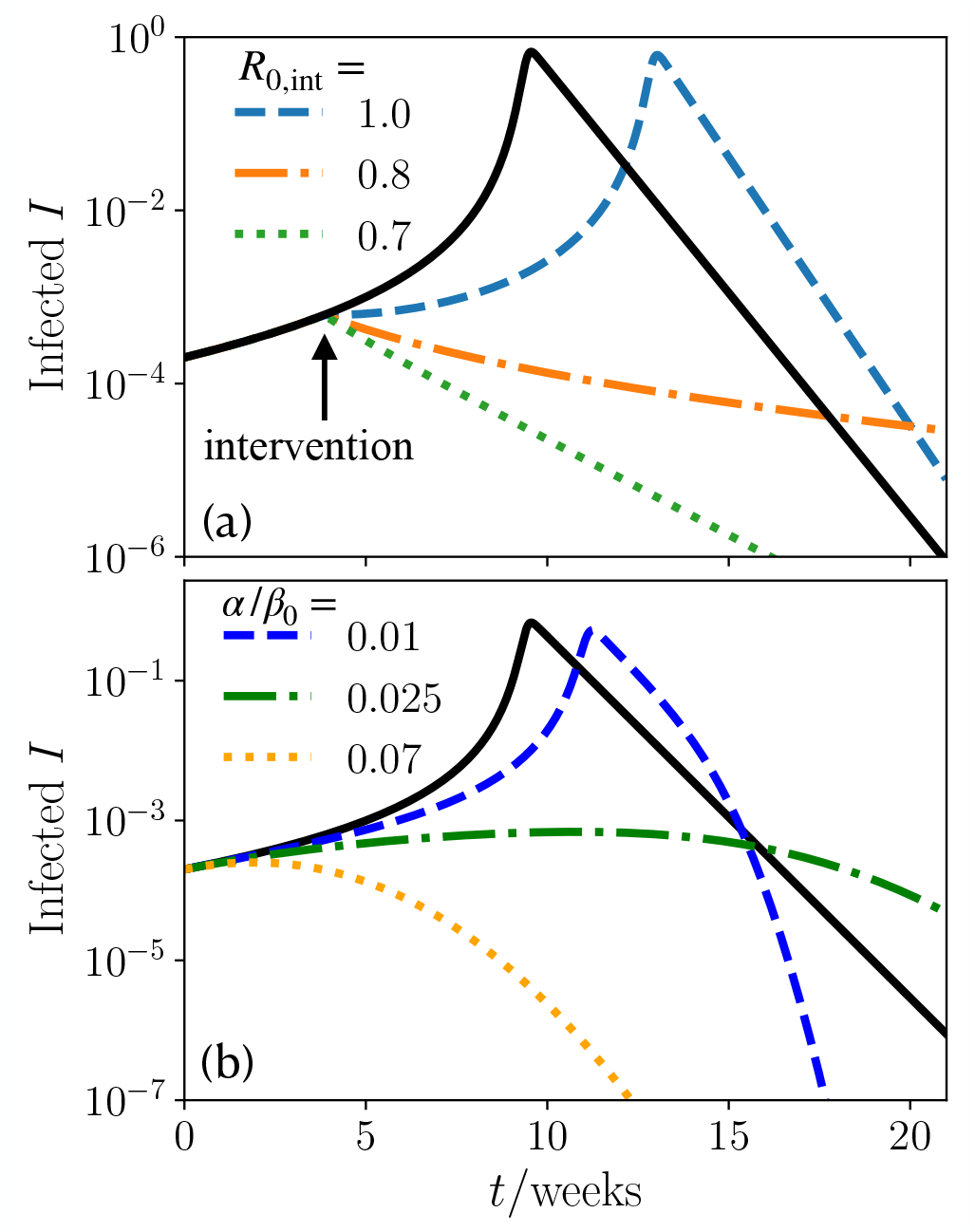
Nonpharmaceutical interventions and vaccinating. (a) Infections as function of time for a super exponential wave. Measures are taken by reducing the reproduction number to *R*_0,int_ at time *t*_int_. (Black line has no intervention. *R*_0_ = 1.2,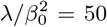, *t*_int_ = 3.85 weeks) (b) Infections as function of time of different vaccination rates *α*. (Black line has no vaccination. *R*_0_ = 1.2, 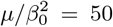, *I*_0_ = 2 × 10^−4^)

In view of the ongoing vaccination campaign against COVID-19 we finally ask how fast vaccination would have to progress in order to suppress a mutation-induced explosion of infection numbers or a mutation-induced rebound at initial reproduction numbers smaller than 1. Vaccinations effectively reduce the number of susceptibles by a vaccination rate *α* (see also Eq. 11). If we start in the explosive (super exponential) regime, a high vaccination rate is needed to interrupt the rapid growth of the infection numbers (Fig. 5(b)): while small vaccination rates (*α/β*_0_ = 0.01) merely shift the infection maximum to a later time, while having little change in the number of infections, larger vaccination rates (*α/β*_0_ = 0.025) effectively suppress the explosion of infection numbers. Therefore, to prevent possible mutation-induced longtime consequences it is imperative to maximize vaccine production worldwide.

## Discussion

Inspired by the ongoing COVID-19 disease and the continued emergence of new mutations supplanting preceding ones, in the present work we have developed a stochastic multistrain generalization of the popular SIR model. Combining this model with coarse graining concepts from statistical physics has allowed us to predict a panorama of possible scenarios for the mutation controlled evolution of infectious diseases.

In particular, our approach suggests that mutations can induce a super-exponential growth of infection numbers in populations which are still from reaching herd-immunity. As compared to the standard exponential growth, interrupting such an super-exponential growth is much more difficult and requires stronger and stronger measures as the disease evolves. In practice, such a super exponential growth may occur e.g. if measures are applied too late, or if vaccines suddenly become ineffective against mutations.

One particularly severe form of such an super exponential growth can occur if the mutation rate of a virus is proportional to the current number of infections. For this case our model predicts a giant infection wave, which is based on a positive feedback loop between the mutation-rate and the infection number causing a massive-self acceleration of the latter resulting in a state where the majority of the population gets infected at the same time. Clearly, such a situation would not only massively overstress any existing health system but once in action it would hardly be interruptable through vaccination.

At later stages of an infectious disease, where the population approaches herd immunity and the infection numbers decrease, an obvious political reaction would be to release measures. However, our simulations suggest that mutations can drive new infection waves even after a longer downwards trend. Such waves can even self-repeat and lead to a pattern of repeated phases of strongly decaying and increasing infection numbers provoking an endless sequence of renewed non-pharmaceutical inventions.

This panorama of mutation induced phenomena which we have identified should inspire detailed modeling works to test them for specific infectious diseases such as COVID-19. These results might also be useful for discussions regarding the importance of a release of vaccine patents to reduce the risk of mutation-induced infection revivals and to coordinate the release of measures following a downwards trend of infection numbers.

## Methods

### Basic reproduction number of COVID-19 mutants

The basic reproduction numbers where extracted from: original variant [46], B.1.1.7 [11], B.1.351 [47], and P.1 [13]. The time point at which a variant has reached 5% in the sequenced genomes reported in [4] (https://nextstrain.org/ncov/global) is used as the emergence time.

### Details on constant mutation rate

We assume that the new infection rates *β*_*n*_ are drawn from a Gaussian distribution, whose mean is the largest current infection rate. Explicitly we have

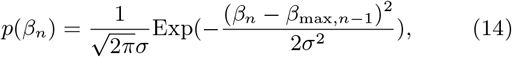

where *β*_max,*n-*1_ denotes the maximal infection rate of the current strains, *β*_*n*_ is the infection rate of the newly mutated strain, *σ* is the standard deviation of the distribution, and new strains are produced at a rate *m*. (In our multi component simulations we use *β*_0_*/γ* = 1, *σ* = 2 × 10^*-*4^, *m/γ* = 2, and the new strain obtains an initial *I*_*n*_(0) = 10^*-*7^)

To coarse grain this mutation model, we assume that the infections immediately assume the maximal infection rate of the newly mutated strain *β*_max,*n*_. To determine *β*_max,*n*_ we compute

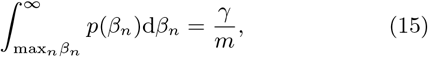

where *m* is the number of times drawn from the distribution Eq. (14). Explicitly, Eq. (15) yields

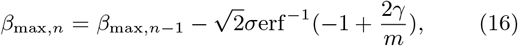

where erf^−1^(*∗*) is the inverse error function and can here be approximated by a negative constant *−C*_1_. We now write the standard deviation as *σ* = *µ*^*∗*^*τ*, with a mutation rate *µ*^*∗*^ and mutation timescale *τ*, giving

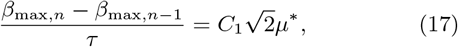

which is a discretized version of 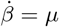, and equivalent to our SIR *β* model (I).

### Details on model beyond constant mutation rate

For model beyond constant mutation rate the infection rates perform a biased random walk. Given an infection rate *β*_*n*_ it will mutate with a probability *p*_0_ and not mutate with probability 1 *- p*_0_. Furthermore, this strain has *I*_*n*_ infections, which are all able to mutate, giving a total mutation probability of 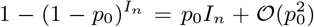. A mutation gives a new strain *I*_*n*_ with an increased *β*_*n*_ = *β*_*n-*1_ + Δ*β*. (In our multi component simulations we use *β*_0_ = 0.1, *γ* = 0.1, Δ*β* = 0.03, *p*_0_ = 2 × 10^−4^, and the new strain obtains an initial *I*_*n*_(0) = 10^−6^)

To coarse grain, we assume that the expectation value of the infection rate ⟨*β*_*n*_⟩ is proportional to the mutation probability. Then a new mutation has the expectation value ⟨*β*_*n*+1_⟩ = *p*_0_*I*_*n*_ and the old strain has ⟨*β*_*n*_⟩ = 1 *− p*_0_*I*_*n*_. Computing the difference gives

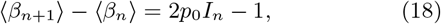

which is a discretization of 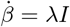, which is our coarse grained infection model (II).

## Data Availability

Data is available upon request.

## Author contributions

H.L. and B.L. designed the research. J.G. and F.J.S. performed the simulations. F.J.S. did the analytical calculations and the simulation data analysis. All authors discussed and wrote the manuscript.

## References

[1] P. Zhou, X.-L. Yang, X.-G. Wang, B. Hu, L. Zhang, W. Zhang, H.-R. Si, Y. Zhu, B. Li, C.-L. Huang, et al., A pneumonia outbreak associated with a new coronavirus of probable bat origin, Nature 579, 270 (2020).

[2] F. Wu, S. Zhao, B. Yu, Y.-M. Chen, W. Wang, Z.-G. Song, Y. Hu, Z.-W. Tao, J.-H. Tian, Y.-Y. Pei, et al., A new coronavirus associated with human respiratory disease in china, Nature 579, 265 (2020).

[3] G. L. Dong E, Du H, An interactive web-based dashboard to track covid-19 in real time., Lancet Inf Dis. 20, 533 (2020).

[4] J. Hadfield, C. Megill, S. M. Bell, J. Huddleston, B. Potter, C. Callender, P. Sagulenko, T. Bedford, and R. A. Neher, Nextstrain: real-time tracking of pathogen evolution, Bioinformatics 34, 4121 (2018).

[5] A. Rambaut, E. C. Holmes, A’. O’Toole, V. Hill, J. T. McCrone, C. Ruis, L. du Plessis, and O. G. Pybus, A dynamic nomenclature proposal for sars-cov-2 lineages to assist genomic epidemiology, Nature Microbiology 5, 1403 (2020).

[6] H. Yao, X. Lu, Q. Chen, K. Xu, Y. Chen, M. Cheng, K. Chen, L. Cheng, T. Weng, D. Shi, et al., Patientderived sars-cov-2 mutations impact viral replication dynamics and infectivity in vitro and with clinical implications in vivo, Cell discovery 6, 76 (2020).

[7] B. Korber, W. M. Fischer, S. Gnanakaran, H. Yoon, J. Theiler, W. Abfalterer, N. Hengartner, E. E. Giorgi, T. Bhattacharya, B. Foley, et al., Tracking changes in sars-cov-2 spike: evidence that d614g increases infectivity of the covid-19 virus, Cell 182, 812 (2020).

[8] N. D. Grubaugh, W. P. Hanage, and A. L. Rasmussen, Making sense of mutation: what d614g means for the covid-19 pandemic remains unclear, Cell 182, 794 (2020).

[9] H. Tegally, E. Wilkinson, M. Giovanetti, A. Iranzadeh, V. Fonseca, J. Giandhari, D. Doolabh, S. Pillay, E. J. San, N. Msomi, et al., Emergence and rapid spread of a new severe acute respiratory syndrome-related coronavirus 2 (sars-cov-2) lineage with multiple spike mutations in south africa, medRxiv (2020).

[10] V. Priesemann, R. Balling, M. M. Brinkmann, S. Ciesek, T. Czypionka, I. Eckerle, G. Giordano, C. Hanson, Z. Hel, P. Hotulainen, et al., An action plan for pan-european defence against new sars-cov-2 variants, The Lancet 397, 469 (2021).

[11] E. Volz, S. Mishra, M. Chand, J. C. Barrett, R. Johnson, L. Geidelberg, W. R. Hinsley, D. J. Laydon, G. Dabrera, A’. O’Toole, et al., Assessing transmissibility of sars-cov-2 lineage b. 1.1. 7 in england, Nature 593, 266 (2021).

[12] N. G. Davies, S. Abbott, R. C. Barnard, C. I. Jarvis, A. J. Kucharski, J. D. Munday, C. A. Pearson, T. W. Russell, D. C. Tully, A. D. Washburne, et al., Estimated transmissibility and impact of sars-cov-2 lineage b. 1.1. 7 in england, Science 372 (2021).

[13] R. M. Coutinho, F. M. D. Marquitti, L. S. Ferreira, M. E. Borges, R. L. P. da Silva, O. Canton, T. P. Portella, S. P. Lyra, C. Franco, A. A. M. da Silva, et al., Model-based evaluation of transmissibility and reinfection for the p. 1 variant of the sars-cov-2, medRxiv (2021).

[14] J. Grauer, H. Löwen, and B. Liebchen, Strategic spatiotemporal vaccine distribution increases the survival rate in an infectious disease like covid-19, Scientific reports 10, 21594 (2020).

[15] S. Zhou, S. Zhou, Z. Zheng, and J. Lu, Optimizing spatial allocation of covid-19 vaccine by agent-based spatiotemporal simulations, GeoHealth , e2021GH000427.

[16] J. Molla, A. P. d. L. Chavez, T. Hiraoka, T. Ala-Nissila, M. Kivelä, and L. Leskelä, Adaptive and optimized covid19 vaccination strategies across geographical regions and age group, arXiv preprint 2105.11562 (2021).

[17] W. O. Kermack and A. G. McKendrick, A contribution to the mathematical theory of epidemics, Proceedings of the royal society of london. Series A, Containing papers of a mathematical and physical character 115, 700 (1927).

[18] H. W. Hethcote, The mathematics of infectious diseases, SIAM review 42, 599 (2000).

[19] H. Andersson and T. Britton, Stochastic epidemic models and their statistical analysis, Vol. 151 (Springer Science & Business Media, 2012).

[20] N. C. Grassly and C. Fraser, Mathematical models of infectious disease transmission, Nature Reviews Microbiology 6, 477 (2008).

[21] J. R. Gog and B. T. Grenfell, Dynamics and selection of many-strain pathogens, Proc. Natl. Acad. Sci. USA 99, 17209 (2002), https://www.pnas.org/content/99/26/17209.full.pdf.

[22] T. Harko, F. S. Lobo, and M. Mak, Exact analytical solutions of the susceptible-infected-recovered (sir) epidemic model and of the sir model with equal death and birth rates, Applied Mathematics and Computation 236, 184 (2014).

[23] M. Kröger and R. Schlickeiser, Analytical solution of the sir-model for the temporal evolution of epidemics. part a: time-independent reproduction factor, J. Phys. A: Math. Theor. 53, 505601 (2020).

[24] R. Schlickeiser and M. Kröger, Analytical solution of the sir-model for the temporal evolution of epidemics: part b. semi-time case, J. Phys. A: Math. Theor. 54, 175601 (2021).

[25] P. Bittihn and R. Golestanian, Stochastic effects on the dynamics of an epidemic due to population subdivision, Chaos 30, 101102 (2020).

[26] S. K. Das, A scaling investigation of pattern in the spread of covid-19: universality in real data and a predictive analytical description, Proc. R. Soc. A. 477, 20200689 (2021).

[27] R. Yaari, G. Katriel, A. Huppert, J. Axelsen, and L. Stone, Modelling seasonal influenza: the role of weather and punctuated antigenic drift, J. R. Soc. Interface. 10, 20130298 (2013).

[28] Y. Zhao, C. Huepe, and P. Romanczuk, Contagion dynamics in self-organized systems of self-propelled agents, arXiv preprint 2103.12618 (2021).

[29] A. Norambuena, F. J. Valencia, and F. Guzm’an-Lastra, Understanding contagion dynamics through microscopic processes in active brownian particles, Scientific Reports 10, 20845 (2020).

[30] B. F. Maier and D. Brockmann, Effective containment explains subexponential growth in recent confirmed covid-19 cases in china, Science 368, 742 (2020).

[31] J. Dehning, J. Zierenberg, F. P. Spitzner, M. Wibral, J. P. Neto, M. Wilczek, and V. Priesemann, Inferring change points in the spread of covid-19 reveals the effectiveness of interventions, Science 369 (2020).

[32] M. Te Vrugt, J. Bickmann, and R. Wittkowski, Effects of social distancing and isolation on epidemic spreading modeled via dynamical density functional theory, Nat. Commun. 11, 5576 (2020).

[33] M. A. Duran-Olivencia and S. Kalliadasis, More than a year after the onset of the covid-19 pandemic in the uk: lessons learned from a minimalistic model capturing essential features including social awareness and policy making, medRxiv (2021).

[34] M. te Vrugt, J. Bickmann, and R. Wittkowski, Containing a pandemic: nonpharmaceutical interventions and the “second wave”, J. Phys. Commun. 5, 055008 (2021).

[35] J. Lasser, J. Sorger, L. Richter, S. Thurner, D. Schmid, and P. Klimek, Assessing the impact of sars-cov-2 prevention measures in schools by means of agent-based simulations calibrated to cluster tracing data, medRxiv (2021).

[36] A. Desvars-Larrive, E. Dervic, N. Haug, T. Niederkrotenthaler, J. Chen, A. Di Natale, J. Lasser, D. S. Gliga, A. Roux, J. Sorger, et al., A structured open dataset of government interventions in response to covid-19, Sci Data 7, 285 (2020).

[37] P. Bittihn, L. Hupe, J. Isensee, and R. Golestanian, Local measures enable covid-19 containment with fewer restrictions due to cooperative effects, EClinicalMedicine 32, 100718 (2021).

[38] X. Zhang, Z. Ruan, M. Zheng, J. Zhou, S. Boccaletti, and B. Barzel, Epidemic spreading under pathogen evolution, arXiv preprint 2102.11066 (2021).

[39] S. Contreras, J. Dehning, M. Loidolt, J. Zierenberg, F. P. Spitzner, J. H. Urrea-Quintero, S. B. Mohr, M. Wilczek, M. Wibral, and V. Priesemann, The challenges of containing sars-cov-2 via test-trace-and-isolate, Nat. Commun. 12, 1 (2021).

[40] E. Estrada, Covid-19 and sars-cov-2. modeling the present, looking at the future, Physics Reports 869, 1 (2020).

[41] C. Yang and J. Wang, A mathematical model for the novel coronavirus epidemic in wuhan, china, Mathematical biosciences and engineering: MBE 17, 2708 (2020).

[42] G. Gonzalez-Parra, D. Mart’inez-Rodr’iguez, and R.J. Villanueva-Mic’o, Impact of a new sars-cov-2 variant on the population: A mathematical modeling approach, Mathematical and Computational Applications 26, 25 (2021).

[43] M. Fudolig and R. Howard, The local stability of a modified multi-strain sir model for emerging viral strains, PloS one 15, e0243408 (2020).

[44] L. Tian, X. Li, F. Qi, Q.-Y. Tang, V. Tang, J. Liu, Z. Li, X. Cheng, X. Li, Y. Shi, et al., Harnessing peak transmission around symptom onset for non-pharmaceutical intervention and containment of the covid-19 pandemic, Nat. Commun. 12, 1 (2021).

[45] D. Meidan, N. Schulmann, R. Cohen, S. Haber, E. Yaniv, R. Sarid, and B. Barzel, Alternating quarantine for sustainable epidemic mitigation, Nat. Commun. 12, 1 (2021).

[46] Y. Liu, A. A. Gayle, A. Wilder-Smith, and J. Rocklöv, The reproductive number of COVID-19 is higher compared to SARS coronavirus, Journal of Travel Medicine 27, 10.1093/jtm/taaa021 (2020).

[47] R. R. Assessment, Risk related to the spread of new sars-cov-2 variants of concern in the eu/eea–first update, European Centre for Disease Prevention and Control An agency of the European Union (2021).

