## Supplementary Information for "Mutation induced infection waves in diseases like COVID-19"

### I. MAXMIMUM OF WAVES IN THE CONSTANT MUTATION RATE MODEL

To obtain some analytical an understanding of the scaling of the maxima in the constant mutation model, we start by considering the equations of susceptible and infections which read

$$\dot{S} = -(\beta_0 + \mu t)SI, \tag{S1}$$

$$\dot{I} = (\beta_0 + \mu t)SI - \gamma I, \tag{S2}$$

$$\tag{S3}$$

where the recovered $R$ follow from $S + I + R = 1$. Redefining the time variable $t' = (\beta_0 + \mu t)$ yields

$$\mu\dot{S} = -t'SI, \tag{S4}$$

$$\mu\dot{I} = t'SI - \gamma I, \tag{S5}$$

$$\tag{S6}$$

where the dot now denotes derivatives with respect to $t'$. Maxima can be found using $\dot{I} = 0$, which is explicity

$$S^* t'^* = \gamma \tag{S7}$$

where $S^*$ are the susceptible at the maximum and $t'^*$ is the time at which the maximum appears. Taking a derivate of $S^*$ with respect to $t'^*$ gives

$$\dot{S}^* = -\frac{\gamma}{(t'^*)^2}, \tag{S8}$$

which is only valid between maxima. Since the susceptible decay monotonically in time, we assume that the $S^*$ also decays monotonically with

$$\dot{S}^* \sim -t'^* I^*, \tag{S9}$$

where $I^*$ are the infected a the maximum. Using Eq. (S8) and Eq (S9) then yields the scaling

$$I^* \sim \frac{\mu}{(\beta + \mu t'^*)^2}. \tag{S10}$$

### II. MATHEMATICAL ANALYSIS OF PHASE DIAGRAM IN BEYOND CONSTANT MUTATION RATE APPROACH

The dynamical equations of our model which goes beyond a constant mutation rate are given by

$$\dot{S} = -\beta SI, \tag{S11}$$

$$\dot{I} = \beta SI - \gamma I, \tag{S12}$$

$$\dot{\beta} = \lambda I, \tag{S13}$$

$$\tag{S14}$$

*

where $R$ follows from $S+I+R=1$. Dividing Eq. (S11)-(S12) by Eq. (S13) gives

$$\frac{dS}{d\beta} = -\frac{\beta S}{\lambda}, \tag{S15}$$

$$\frac{dI}{d\beta} = \frac{\beta S}{\lambda} - \frac{\gamma}{\lambda}, \tag{S16}$$

where $\beta$ reparameterises time. Equation (S15) is solved by

$$S = S_0 e^{\frac{\beta_0^2-\beta^2}{2\lambda}}, \tag{S17}$$

with initial susceptible $S_0$ and initial infection rate $\beta_0$. Equation (S16) can be rewritten as

$$dI = \frac{\beta S}{\lambda} d\beta - \frac{\gamma}{\lambda} d\beta \tag{S18}$$

$$= -dS - \frac{\gamma}{\lambda} d\beta, \tag{S19}$$

where we used Eq. (S15). Integrating then yields

$$I - I_0 = S_0 - S - \frac{\gamma}{\lambda}(\beta - \beta_0) \tag{S20}$$

To obtain the phase diagram shown in the main text, we need to look for the extrema of the infections. Extrema of $I$ are given by the condition $\dot{I}=0$ which explicitly read

$$S^*\beta^* - \gamma = 0, \tag{S21}$$

where $S^*$ are the susceptible at the extremum and $\beta^*$ is the infection rate at the extremum. Using Eq. (S17) gives

$$\frac{\gamma}{\beta^*} = S_0 e^{\frac{\beta_0^2-(\beta^*)^2}{2\lambda}}, \tag{S22}$$

which is solved by

$$\beta^* = -i\sqrt{\lambda}\left(W\left(-\frac{\gamma^2}{\lambda S_0^2} e^{-\frac{\beta_0^2}{\lambda}}\right)\right)^{1/2} \tag{S23}$$

where $W(*)$ is the Lambert function. The argument in the Lambert function of Eq (S23) is smaller than zero. This gives rise to two solutions, implying two extrema of the infections. Explicitly the extrema $I^*$ can be found by using

$$I^* = I_0 + S_0 - \frac{\gamma}{\beta^*} - \frac{\gamma}{\lambda}(\beta^* - \beta_0), \tag{S24}$$

together with Eq (S23). Furtermore, to determine maxima and minima we can use

$$\ddot{I}|_{S^*\beta^*=\gamma} = (I^*)^2 S^*(\lambda - (\beta^*)^2) \tag{S25}$$

such that the sign of $(\lambda - (\beta^*)^2)$ decides between minimum and maximum.

### III. CRITICAL SHORT TIME BEHAVIOR IN BEYOND CONSTANT MUTATION RATE APPROACH

At small times we can approximate the number of susceptible people by $S \approx 1$, which gives the set of equations

$$\dot{I} = \beta[I]I - \gamma I, \tag{S26}$$

$$\dot{\beta} = \lambda I, \tag{S27}$$

with the initial condition $\beta(0) = \beta_0$. By changing the initial condition of Eq.(S27) to $\beta(0) = \beta_0 - \gamma$ we can rewrite Eq.(S26) as

$$\frac{d\ln I}{dt} = S_0\beta[I]. \tag{S28}$$

We now use the transformation $\omega = \ln I$, giving

$$\frac{d\omega}{dt} = S_0\beta\left[e^{\omega}\right] \tag{S29}$$

$$\dot{\beta} = \lambda e^{\omega}. \tag{S30}$$

Taking a derivatite of Eq.(S29), gives

$$\frac{d^2\omega}{dt^2} = S_0\dot{\beta} = S_0\lambda e^{\omega}, \tag{S31}$$

which has initial conditions $\omega(0) = \ln(I(0))$ and $\dot{\omega}(0) = \beta - \gamma$ and can be solved explicitly. Putting back the transformation, we arrived at the following expression for the number of infected people

$$I(t) = -\frac{\delta_1}{\lambda\left[1 + \cosh\left(2\ln\delta_2 + t\sqrt{\delta_1}\right)\right]}, \tag{S32}$$

with $\delta_1 = -2I_0\lambda + (\gamma - S_0\beta_0)^2$ and $\delta_2 = -\frac{\sqrt{\gamma - S_0\beta_0 + \sqrt{\delta_1}}}{\sqrt{-\gamma + S_0\beta_0 + \sqrt{\delta_1}}}$. Equation (S32) has poles at

$$t_c = \frac{-2\log(\delta_2) + i\pi(2k-1)}{\sqrt{\delta_1}}, \tag{S33}$$

where $k$ is an integer and the physically relevant critical time $t_c$ is given by $k$ yielding the smallest positive $t_c$. Expanding Eq. (S32) around the critical time $t_c$ gives

$$I(t) = \frac{2}{\lambda}\frac{1}{(t-t_c)^2}. \tag{S34}$$

### IV. INDEPENDENT PARAMETERS OF COARSE GRAINED MSIR MODEL

The equations of our constant mutation rate coarse grained model are

$$\dot{S} = -\beta S I, \tag{S35}$$

$$\dot{I} = \beta S I - \gamma I, \tag{S36}$$

$$\dot{\beta} = \mu. \tag{S37}$$

From the condition $S + I + R = 1$ it then follows that $R(t=0) = 1 - I_0 - S_0$, such that we have three parameters $I_0$, $S_0$ and $\beta_0$ for the initial condition. Additionally, we have the parameters $\mu$ and $\gamma$, making our parameter space five dimensional.

We continue considering the units of each parameter and variable which are $[S] = 1$, $[I] = 1$, $[\beta] = 1/\text{time}$, $[\beta_0] = 1/\text{time}$, $[\gamma] = 1/\text{time}$ and $[\mu] = 1/\text{time}^2$. By nondimensionalizing Eq.(S35)-(S37) we find

$$\dot{\mathcal{S}} = -\beta\mathcal{S}\mathcal{I}, \tag{S38}$$

$$\dot{\mathcal{I}} = \beta\mathcal{S}\mathcal{I} - \mathcal{I}, \tag{S39}$$

$$\dot{\beta} = \mu/\beta_0^2. \tag{S40}$$

with $\mathcal{S} = S\beta_0/\gamma$, and $\mathcal{I} = I\beta_0/\gamma$. The parameter space then reduces to three dimensions where the nondimensional parameters are the initial infected $I_0$, the basic reproduction number $S_0\beta_0/\gamma$ and the mutation rate $\mu/\beta_0^2$. The calculation for our model that goes beyond a constant mutation rate is analogous, with the resulting nondimensional mutation rate $\lambda/\beta_0^2$.
